# Efficacy of “stay-at-home” policy and transmission of COVID-19 in Toronto, Canada: a mathematical modeling study

**DOI:** 10.1101/2020.10.19.20181057

**Authors:** Pei Yuan, Juan Li, Elena Aruffo, Qi Li, Tingting Zheng, Nick Ogden, Beate Sander, Jane Heffernan, Evgenia Gatov, Effie Gournis, Sarah Collier, Yi Tan, Jun Li, Julien Arino, Jacques Bélair, James Watmough, Jude Dzevela Kong, Iain Moyles, Huaiping Zhu

## Abstract

**Background:** In many parts of the world, restrictive non-pharmaceutical interventions (NPI) that aim to reduce contact rates, including stay-at-home orders, limitations on gatherings, and closure of public places, are being lifted, with the possibility that the epidemic resurges if alternative measures are not strong enough. Here we aim to capture the combination of use of NPI’s and reopening measures which will prevent an infection rebound.

**Methods:** We employ an SEAIR model with household structure able to capture the stay-at-home policy (SAHP). To reflect the changes in the SAHP over the course of the epidemic, we vary the SAHP compliance rate, assuming that the time to compliance of all the people requested to stay-at-home follows a Gamma distribution. Using confirmed case data for the City of Toronto, we evaluate basic and instantaneous reproduction numbers and simulate how the average household size, the stay-at-home rate, the efficiency and duration of SAHP implementation, affect the outbreak trajectory.

**Findings:** The estimated basic reproduction number R_0 was 2.36 (95% CI: 2.28, 2.45) in Toronto. After the implementation of the SAHP, the contact rate outside the household fell by 39%. When people properly respect the SAHP, the outbreak can be quickly controlled, but extending its duration beyond two months (65 days) had little effect. Our findings also suggest that to avoid a large rebound of the epidemic, the average number of contacts per person per day should be kept below nine. This study suggests that fully reopening schools, offices, and other activities, is possible if the use of other NPIs is strictly adhered to.

**Interpretation:** Our model confirmed that the SAHP implemented in Toronto had a great impact in controlling the spread of COVID-19. Given the lifting of restrictive NPIs, we estimated the thresholds values of maximum number of contacts, probability of transmission and testing needed to ensure that the reopening will be safe, i.e. maintaining an *R*_*t*_ < 1.

**Research in context:** *Evidence before this study:* A survey on published articles was made through PubMed and Google Scholar searches. The search was conducted from March 1 to August 13, 2020 and all papers published until the end of this research were considered. The following terms were used to screen articles on mathematical models: “household structure”, “epidemic model”, “SARS-CoV-2”, “COVID-19”, “household SIR epidemic”, “household SIS epidemic”, “household SEIR epidemic”, “quarantine, isolation model”, “quarantine model dynamics”, “structured model isolation”. Any article showing, in the title, application of epidemic models in a specific country/region or infectious diseases rather than SARS-CoV-2 were excluded. Articles in English were considered.

*Added value of this study:* We develop an epidemic model with household structure to study the effects of SAHP on the infection within households and transmission of COVID-19 in Toronto. The complex model provides interesting insights into the effectiveness of SAHP, if the average number of individuals in a household changes. We found that the SAHP might not be adequate if the size of households is relatively large. We also introduce a new quantity called symptomatic diagnosis’ completion ratio (d_c). This indicator is defined as the ratio of cumulative reported cases and the cumulative cases by episode date at time t, and it is used in the model to inform the implementation of SAHP. If cases are diagnosed at the time of symptom onset, isolation will be enforced immediately. A delay in detecting cases will lead to a delay in isolation, with subsequent increase in the transmission of the infection. Comparing different scenarios (before and after reopening phases), we were able to identify thresholds of these factors which mainly affect the spread of the infection: the number of daily tests, average number of contacts per individual, and probability of transmission of the virus. Our results show that if any of the three above mentioned factors is reduced, then the other two need to be adjusted to keep a reproduction number below 1. Lifting restrictive closures will require the average number of contacts a person has each day to be less than pre-COVID-19, and a high rate of case detection and tracing of contacts. The thresholds found will inform public health decisions on reopening.

*Implications of all the available evidence:* Our findings provide important information for policymakers when planning the full reopening phase. Our results confirm that prompt implementation of SAHP was crucial in reducing the spread of COVID-19. Also, based on our analyses, we propose public health alternatives to consider in view of a full reopening. For example, for different post-reopening scenarios, the average number of contacts per person needs to be reduced if the symptomatic diagnosis’ completion ratio is low and the probability of transmission increases. Namely, if fewer tests are completed and the usage of NPI’s decreases, then the epidemic can be controlled only if individuals can maintain contact with a maximum average number of 4-5 people per person per day. Different recommendations can be provided by relaxing/strengthening one of the above-mentioned factors.

## 1. Introduction

COVID-19 has been spreading rapidly across the world since the first outbreak in late 2019 in Wuhan, China, and severely affecting economies and health systems globally. On March 11, 2020, the World Health Organization (WHO) classified the disease as a global pandemic and the cases have been increasing daily since then^1^.

In the absence of specific antivirals and vaccines, COVID-19 can be mitigated via non-pharmaceutical interventions (NPIs), i.e., social distancing (including SAHP), isolation of cases, contact tracing, quarantine, as well as personal protection methods of hand washing, and wearing of masks or other personal protective equipment (PPE). NPIs have been shown to be effective in mitigating COVID-19 spread^2,3,4,5^. For the purpose of effective control, the Canadian Government has strongly encouraged residents to take any possible precautions to protect themselves^6^, while Provinces and Territories have implemented restrictive closures of businesses, schools, work and public spaces, to reduce the number of contacts among people. Ontario declared a state of emergency on March 17. Since then, the City of Toronto has issued directions on a series of NPIs^7^.

The sharp increase of COVD-19 infectious cases can overload the healthcare system. The “stay-at-home” policy has deeply modified daily routines, reducing contacts outside the household, but also possibly increasing contacts with family members, which can lead to higher transmission risk within a household^8^, where the secondary infection rate in household contacts can be as high as 30%^9^. However, even with this increased risk at home, the SAHP may be beneficial for control in the community^10^. Different studies investigated the transmission within households^9,11,12^. Keeling^12^ extended an SIR model where two transmission regimes are considered (within and outside household). These studies have all shown the importance of within and between household transmission, however, for infections requiring a household quarantine and SAHP, it is fundamental to consider the period of time that individuals spend in the household. In fact, as more people are following SAHP and isolation, the intra-family contact relations will change, consequently affecting the probability of transmission among family members.

If everyone stays at home, the contact rate will be greatly decreased, which will quickly reduce the infection rate and, thus, control the epidemic. However, this is not practically possible. The essential operation of society still needs people to continue working, and to contact others in the process of obtaining essentials such as groceries. Moreover, when the duration of SAHP is too long, it has negative impact on individuals’ physical and mental health as well as on the economy^13,14^. So, restrictive closures need to be lifted as soon as possible. Due to different attitudes towards the epidemic, the speed of compliance to NPIs will vary. Therefore, the rate at which people “stay-at-home” is a function of changes in policies and behaviors over time. Also, the stay-at-home rates for symptomatic cases, or for traced contacts, are different from that of uninfected or asymptomatic individuals, since there will be some form of compulsory home isolation/quarantine after diagnosis (for cases) or tracing of contacts^15^. Rates of diagnosis and isolation of cases, and tracing and quarantine of contacts, as well as public compliance to SAHP will be important factors determining rates of transmission and likelihood of epidemic resurgence after lifting of restrictive closures^31^.

To allow for this level of complexity, we developed a household-based transmission model that further captures differences in policy uptake behaviors. We aimed to evaluate the effect of SAHP on the transmission of COVID-19, accounting for average household size, the rates with which people respond and comply with the policy, as well as the length of the policy implementation. Additionally, based on the average family size of Toronto and epidemic data, we computed the reproduction numbers *R*_0_ and *R*_*t*_. We also investigated the conditions on the number of contacts, testing, and use of NPIs which will maintain an effective reproduction number below 1 as well as simulate the dynamic behavior under different reopening scenarios, assuming that SAHP has been relaxed. Our simulations propose reopening strategies to public health.

## 2. Method

### 2.1 Data and materials

According to the 2016 census, 2,731,571 people live in the city of Toronto, representing 21% of the population of Ontario, and the average household size in the city is 2.4^16^. Like other major cities in Canada, Toronto experienced a large number of COVID-19 cases. We obtained daily new confirmed cases data, by episode date and reporting date in Toronto from Feb 24, 2020, to Jun 27, 2020. (see Figure 1A)^15,17^. Due to the lack of hospital resources, testing reagents, and the waiting time for testing, there is a time lag between the episode date and the reporting date (Figure 1A). We have chosen to use data by episode date, which is accepted to be more in line with the real epidemic situation. Based on these data and available case information, it is apparent that earlier cases were imported, so we ignore data prior to Feb 24. We use data only until Jun 13 to fit the model, which is two weeks before Jun 27 (period due to the incubation time plus the reporting delay), to ensure minimum error.

**Figure 1.**
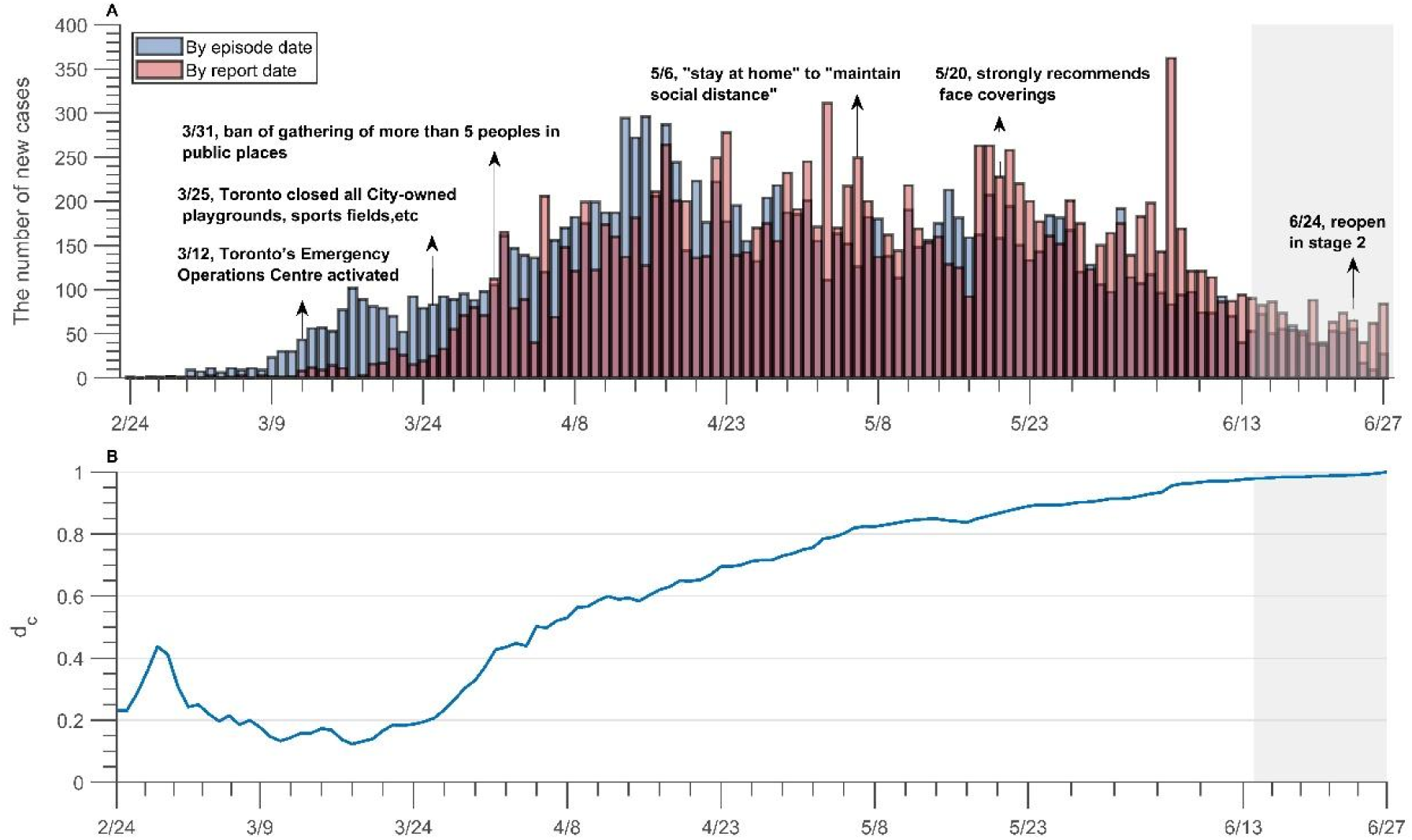
COVID-19 cases in Toronto by report date and episode date. (A)The daily new case of infection by episode date and first report date. (B) The change of *d*_*c*_ over time for the city of Toronto from Feb. 24 to June 27, 2020. *d*_*c*_ =symptomatic diagnosis’ completion ratio.

In Toronto, testing has mainly been provided to individuals showing symptoms^15^. Using the reporting and episode date data, we define

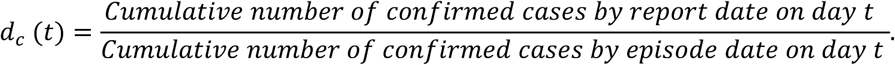

as an indicator with *d*_*c*_(*t*) ∈ [0,1], the ratio of symptomatic diagnosis’ completion. This quantity is used to inform the stay-at-home rate of detected infectious people (i.e. the rate at which they follow isolation recommendations). It is obvious that a delay in case diagnosis will result in a delay in implementing control measures, increasing the risk of transmission. Alternatively, a quick diagnosis of symptomatic cases will lead to a faster implementation of SAHP. Due to the limitation of the data length used in this study, *d*_*c*_(*t*) is unknown after Jun 13, and we will assume that it is maintained at the level of June 13 in the simulations.

### 2.2 Compartmental model: description and assumptions

We develop a household-based transmission model following a Susceptible-Exposed-Asymptomatic (subclinical) – Infectious (prodromal phase)-Infectious (with symptoms) framework including two further compartments, depending on the severity of the infection: hospitalization (H) and fully isolated (W). Given the importance of asymptomatic and pre-symptomatic infection in COVID-19^18^, both stages are included. To capture differences in social policy uptake, we further divide the population in two subgroups: individuals following SAHP (be that associated with recommendations for all citizens to stay at home, or associated with orders to isolate at home for mild cases, and for at-home quarantine of contacts), and those not following it. Based on the process of SAHP implementation, we assumed that the time needed to complete it follows a Gamma distribution.

The flow diagram in Figure 2 describes the dynamics of our model. Tables 1-3 report the assumptions, variables and parameters employed in the study, respectively. Details on stay-at-home, quarantine and isolation ratios (see below), household structure and model equations are provided in Appendix A.

**Table 1.** Model assumptions.

|  |  |  |
| --- | --- | --- |
| General<br>setting | a | No birth, death or immigration. |
| | b | We divide the population into two groups: one consisting of individuals who follows SAHP (marked by subscript $q$) and another consisting of individuals who do not opt for this intervention (marked by subscript $g$). Due to influences of self-protection consciousness and severity of the epidemic, people are assumed to move from one group to another with stay-at-home rate (denoted by $q(t)$) or going out rate (denoted by $g(t)$). |
| | c | Each subpopulation is further the divided into Susceptible ($S_i(t)$), Exposed ($E_i(t)$), Asymptomatic (subclinical) infection ($A_i(t)$), Infectious pre-symptomatic (will eventually show symptoms) ($I_{i1}(t)$) and Infectious symptomatic ($I_{i2}(t)$). |
| | d | Both $A_i(t)$ and $I_{i1}(t)$ are infectious virus carriers. Individuals in $A_i(t)$ will never show symptoms, while individuals in $I_{i1}(t)$ develop into symptomatic classes ($I_{i2}(t)$) after a specified period of time. |
| | e | Mild symptomatic infections ($I_{i2}(t)$), may choose to either isolate themselves at home (or other places). If the quarantine is respected well enough, these infections will be fully isolated and, consequently, will not contribute to the spread of the virus. Otherwise, they are still a source of infection until recovery. |
| | f | Two further compartments for severe infections: the fully isolated ($W(t)$), and the hospitalized ($H(t)$) who are all severely affected. Neither of these compartments contribute to infection transmission. |
| Household<br>structure<br>setting | g | All households contain $n$ ($n = 3$) individuals and family members are homogeneously mixing i.e, contacting each other randomly. |
|  | h | The infection rate of the asymptomatic and symptomatic infectious individuals to the susceptible is the same among the household. |
| | i | Two members in a family cannot be infected at the same time $t$. |
|  | j | Every family except for those with symptomatic members has an equal opportunity to be released from quarantine after the SAHP is relaxed. |
|  | k | Households with infected symptomatic individuals will continue to be quarantined after the SAHP is relaxed. |
| | l | For family members following SAHP, susceptible $S_q(t)$ will only be infected by infectious individuals in the home $A_q(t)$, $I_{q1}(t)$ or $I_{q2}(t)$. |
|  | m | When no infections in a household, the family will be safe and will no longer be involved in the transmission of COVID-19. |

Note: See appendix for model details and derivation process

**Table 2.** Identification of the variables and their initial values.

| **Variable**<br>**s** | **Descriptions** | **Fixed initial**<br>**Values** | **Sources** |
| --- | --- | --- | --- |
| $I_{g2}(t)$ | The number of SAHP non-compliant infected individuals with symptoms. | **10** | Data |
| $S_q(t)$ | The number of SAHP compliant susceptible individuals. | $(n-1)*3$ | Calculate<br>d |
| $E_q(t)$ | The number of SAHP compliant exposed individuals | 0 | |
| $A_q(t)$ | The number of SAHP compliant inapparent (subclinical) infected individuals | 0 | |
| $I_{q1}(t)$ | The number of SAHP compliant infected individuals that do not symptoms that will become symptomatic | 0 | |
| $I_{q2}(t)$ | The number of SAHP compliant infected with symptoms. | 3 | **17** |
| $H(t)$ | The number of patients in hospitals | 0 | |
| $W(t)$ | The number of isolation patients | 0 | |
| $P$ | Total number of populations in Toronto | 2956024 | **27** |
| **Variable**<br>**s** | **Descriptions** | **Initial Values** | **Sources** |
| $S_g(t)$ | The number of SAHP non-compliant susceptible individuals | 2955988 | Estimated |
| $E_g(t)$ | The number of SAHP non-compliant exposed individuals | 20 | Estimated |
| $A_g(t)$ | The number of SAHP non-compliant inapparent infected at day t. (that will never develop symptoms) | 1 | Estimated |
| $I_{g1}(t)$ | The number of SAHP non-compliant infected without symptoms at day t. (that will become symptomatic) | 2 | Estimated |

**Table 3.** Parameter estimation for COVID-2019 in Toronto [a, Wuhan/Toronto].

| Parameter<br>s | Descriptions | Fixed Values | Sources |
| --- | --- | --- | --- |
| $\tau_1$ | Average time spent in the exposed classes, $E_g, E_q$, days | 4 | 23,28 |
| $\tau_2$ | Average time period spent in $I_{g1}, I_{q1}$, days | 3 | 28 |
| $a$ | Proportion of infected with apparent infection | 0.953 | 3 |
| $\gamma_a$ | Recovery rate of inapparent infected | 0.07 | 3 |
| $\gamma_m$ | Recovery rate of patients with mild symptoms | 1/14 | 1 |
| $\gamma$ | Recovery rate of patients in hospitals | 0.0357 | 1(1/42-1/21) |
| $c_0$ | Contact rate before SAHP implemented, 1/day | 11.58 | 29 |
| $T_1$ | Time when the SAHP is implemented | March 12 | 7 |
| $T_2$ | Time when SAHP is relaxed | May 6 | 7 |
| $T_3$ | Time when the reopening of stage 1 begins | May 19 | 30 |
| $T_4$ | Time of reopening of stage 2 begins | June 24 | 7 |
| $n$ | Average number of household population | 2-3 | 16 |
| $q(t)$ | Stay-at-home rate of $S_g, E_g, A_g$ and $I_{g1}$ | - | |
| $d_c(t)$ | Completion rate of diagnosis of all symptomatic infections | - | 15 |
| $g(t)$ | Proportion of households of going out after relaxing the SAHP | - | |
| $q_{g2}(t)$ | Quarantined rate of $I_{g2}$ | - | |
| $Q(t)$ | The proportion of population in stay-at-home state to the total population at time t | - | |
| Parameter<br>s | Descriptions | Estimated<br>Values | Sources |
| $\beta_g$ | Probability of transmission per contact outside household | 3.2984e-02 | Estimated |
| $\mu$ | Exponential decreasing rate of contact rate due to SAHP | 7.5000e-01 | |
| $\beta_q$ | Infection rate of SAHP compliant susceptible by the infectious one without symptoms | 1.5030e-02 | Estimated |
| $q$ | Stay-at-home rate of $S_g, E_g, A_g$ and $I_{g1}$ before SAHP implemented | 3.0001e-04 | Estimated |
| $\varepsilon$ | Adjust parameter | 7.0000e-01 | Estimated |
| $g$ | Going out rate of $S_g, E_g, A_g$ and $I_{g1}$ during the period of SAHP implemented | 1.0000e-04 | Estimated |
| $\theta_h$ | Proportion of hospitalization of $I_{g2}$ | 0.0152 | Estimated |
| $\theta_i$ | Isolation rate of confirmed cases | 3.9978e-02 | Estimated |
| $d$ | Disease-induced death rate in hospitals | 3.4000e-02 | Estimated |
| $Q$ | Maximum compliance rate induced by SAHP | 6.5058e-01 | Estimated |
| $\Delta T_Q$ | Average completing time for all those who conducted the SAHP | 9 | Estimated |
| $G_0$ | Maximum going out rate in the period of May 6 to May 19 | 1.5000e-01 | Estimated |
| $G_1$ | Maximum going out rate in the period of May 20 to Jun 24 | 3.0000e-01 | Estimated |
| $G_2$ | Maximum going out rate in the period of reopen stage 2 starting Jun 24 | 3.0000e-01 | Assumed |
| $\Delta T_G$ | Average time required for all stay-at-home people to go out after reopening | 3 | Assumed |

**Figure 2.**
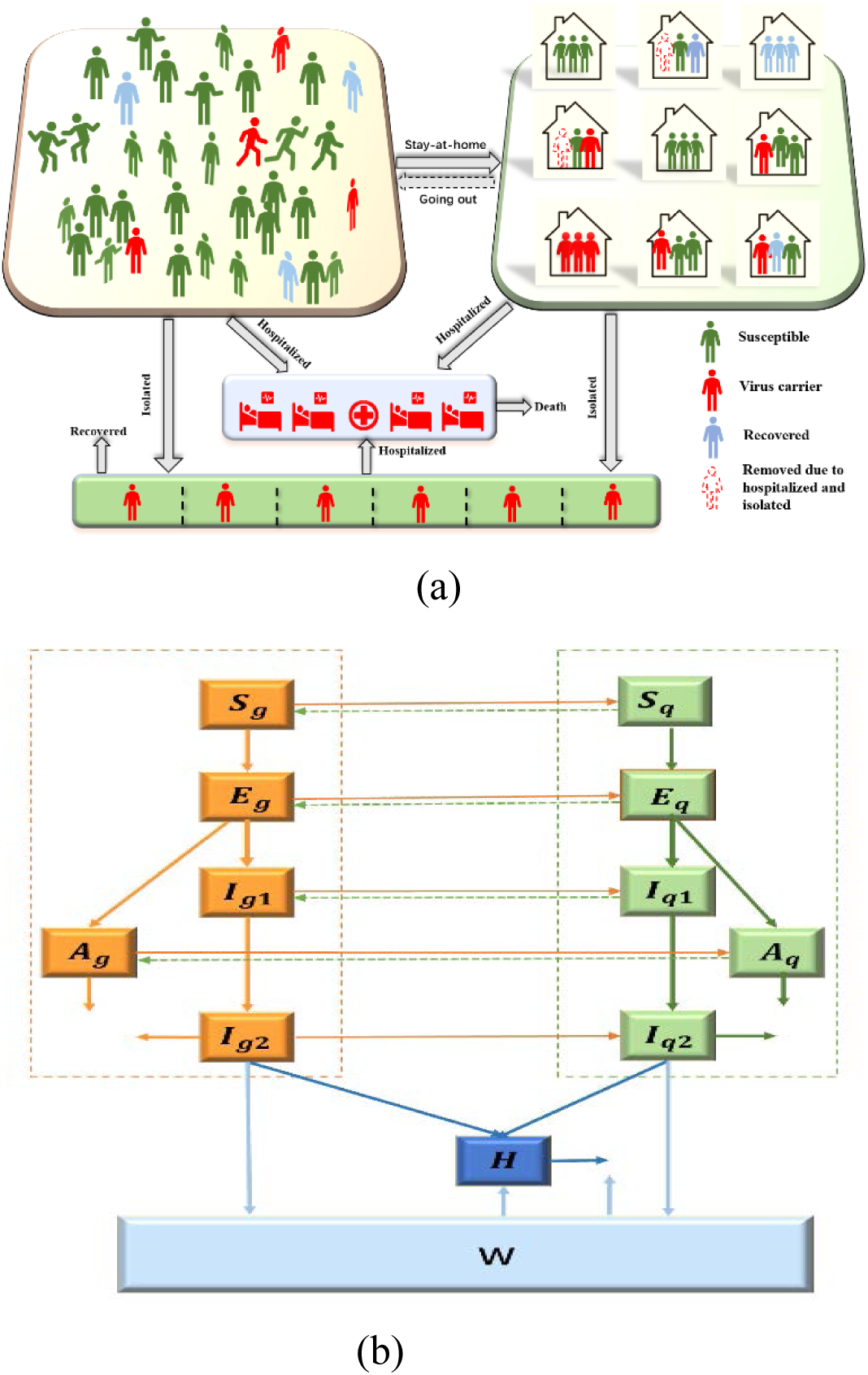
Modeling with household structure. (a) shows the activity and response of different groups. (b) Schematic diagram of the dynamics of COVID-19 in Toronto. Solid lines indicate movement between classes. Dashed lines represent the virus transmission routes.

The model is structured over two time periods: before and after the implementation of SAHP as a population-wide NPI. In the latter case, the population is divided into SAHP compliant and non-compliant subpopulations.

The movement between the SAHP compliant and non-compliant groups is modeled as policy and time vary, described by a stay-at-home rate *q*(*t*) and a going out rate *g*(*t*). Before the city of Toronto declared state of emergency on March 12^7^, due to the impact of self-prevention awareness and the severity of the epidemic, a small number of people would consciously stay at home. We, therefore include a small stay-at-home rate in the absence of governmental SAHP policy. After measures were implemented, some people chose to stay at home based on their own behaviors and their knowledge of the epidemic. We assume that τ is a random variable which describes how long it will take the five groups *S*_*g*_, *E*_*g*_, *A*_*g*_, *I*_*g*1_, *I*_*g*2_ to complete the stay-at-home process when conducting SAHP. Although *I*_*g*2_ is the symptomatic compartment, until confirmed, we assume that its stay-at-home rate is the same as the others *g* groups.

As well as the SAHP, other NPIs in operation include detection and isolation of COVID-19 cases by testing, and tracing and quarantine of people contacting detected cases. Here these other NPIs are modelled together simply as isolation of cases if they are serious enough to be hospitalized, and stay-at-home rates for infectious people that are detected and mild, as well as infectious people who were contacts with cases, were traced and placed in quarantine. The “quarantine” rate of *I*_*g*2_ is essentially a stay-at-home rate that is higher than that of the general population and is defined as *q*_*g*2_(*t*) = *q*(*t*) + *εd*_*c*_(*t*). If the testing process is not included, *q*_*g*2_(*t*) = *q*(*t*).

### 2.3 Reproduction numbers

#### Model-free estimation of the reproduction number

The basic reproduction number *R*_0_ is numerically estimated using an exponential growth method^19,20^based on the Toronto case data by episode date^15,17^. The instantaneous reproduction number *R*_*t*_ is also estimated by Wallinga and Teunis-type approach^21,22^. *R*_*t*_ is defined as

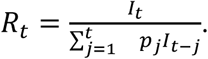

where *I*_*t*_ is the new cases on day *t* and *p*_*j*_ is the discretized distribution of serial interval, assuming a Gamma distributed serial interval of 7.5 days with standard deviation of 3.4 days^23^.

#### Model-based estimation of reproduction number

Although informative, the previous *R*_*t*_ is evaluated only on symptomatic cases, and can thus can underestimate *R*_0_. Hence, we used total infection data (including symptomatic and asymptomatic infection) generated by the model to estimate the instantaneous reproduction number *R*_*t*_ in Toronto.

#### Risk index after reopening

We define a risk index *R*_*reopen*_ to evaluate the risk of reopening by calculating the reproduction number without SAHP:

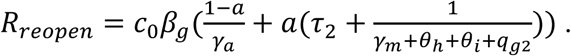

## 3. Results

### 3.1 Parameter estimation and data fitting

Using the cumulative confirmed case data by episode date and the cumulative number of deaths data in Toronto from Feb 24 to Jun 13, we fit our model by the least-square method to estimate the parameters. The results show that our model fits very well with the Normalized Mean Square Error (NMSE) = 0.998(Figure 3A).

**Figure 3:**
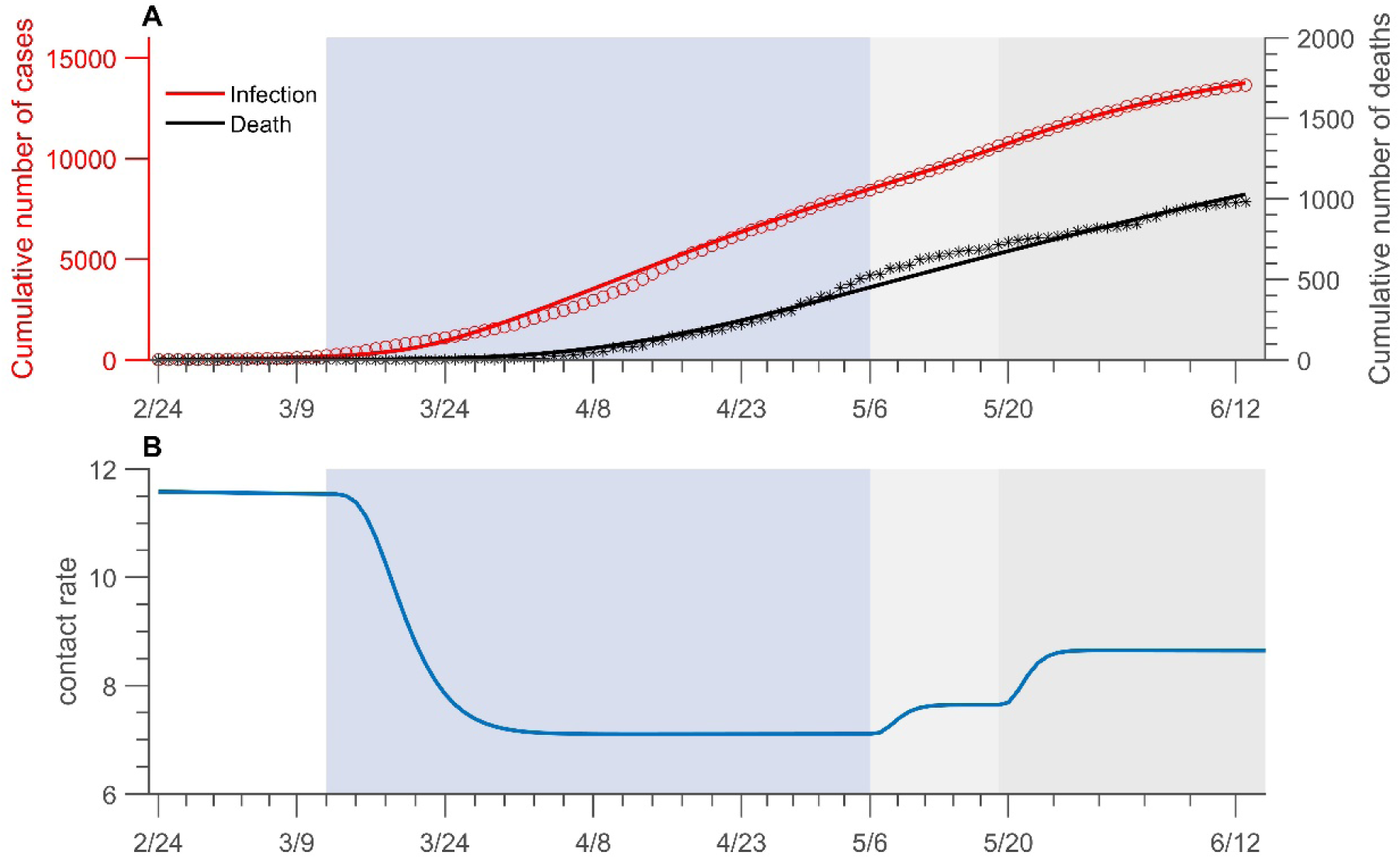
Cumulative COVID-19 incidence and deaths in Toronto (A) and the change of contact rate over time. Data fitting of COVID-19 infection in Toronto from Feb 24 to Jun 13, 2020. The red circles (infection) and black stars (death, right panel) represent real data. The solid curves are from model simulations. Shaded bars show the dates that SAHP implemented, (light blue), preopening (light grey), reopening stage 1 (medium grey). All dates are in 2020.

The results of parameter estimation indicate that at most 65.1% of people stay at home due to SAHP, after which the contact rate dropped from an initial 11.58 to 7.1, with a reduction of 39%. After May 6, it increased to 8.65, and after the stage 1 reopening of the city on May 19^7^, it gradually increased to 9.4, corresponding to an 18% and 24% increase compared to May 6, respectively (Figure 3B).

### 3.2 Estimation of reproduction numbers in Toronto

The estimation result of the model-free *R*_*c*_ is 1.45 (95% CI 1.43-1.48) (goodness of fit *R*^2^ = 0.905), while the model-based *R*_*c*_ is 2.36 (2.28-2.45) (*R*^2^ = 0.971). According to the episode data, *R*_*t*_ varied before and after the implementation of SAHP, which gradually decreased from 3.56 (95% CI 3.02-4.14) on March 12 to less than 1 on April 22 and to 0.84 (0.79-0.89) on May 6, corresponding to a 76% (71-81%) reduction in transmissibility (Figure 4A). After May 6, launching ActiveTO plan^15^, *R*_*t*_ gradually increased and surpassed 1, rising to 1.13 (1.07-1.20) on May 19^7^. The increase in transmissibility associated with ActiveTO plan was 26% (17-34%) (Figure 4B). But after entering the first phase of the city restart on May 19^7^, *R*_*t*_ showed a clear downward trend, and gradually decreased to 0.67 (0.61-0.73) on June 13, although the contact rate was expected to be higher. The probability of transmission per contact after May 19 would be lower than before.

**Figure 4:**
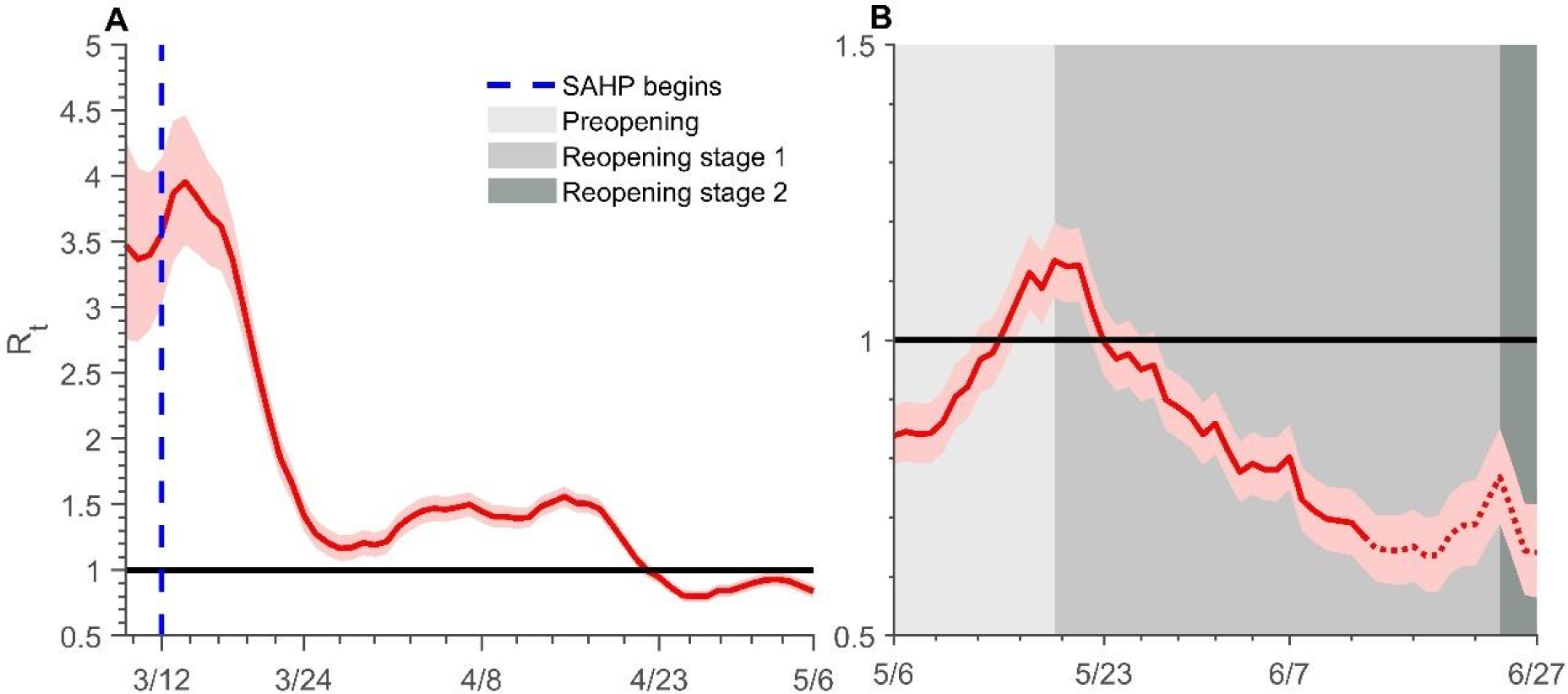
Transmissibility of COVID-19 in Toronto. Estimates of daily *R*_*t*_ of COVID-19 over time (A) from Mar 8 to May 6 and (B) from May 6 to Jun 27, with 95% CIs represented by the pink shaded area. The dates after Jun 13 are indicated by a red dotted line. The dark solid line indicates the critical threshold of *R*_*t*_ = 1. The blue dashed line is the time that the SAHP activated. Shaded bars show the dates of preopening (light grey), reopening stage 1 (medium grey) and stage 2 (dark grey). All dates are in 2020. *R*_*t*_= instantaneous reproduction number.

### 3.3 Effect of stay-at-home policy

By implementing SAHP up to May 6, the cumulative number of infections dropped significantly compared to without SAHP (Figure 5A, B). The aggregate number of infected persons without SAHP was 12.5 times that of conducting SAHP with a mean family size (*n*) of 3. Moreover, when *n* is smaller, the effect of SAHP on the control of the epidemic is better. The cumulative number of infected people on May 6 with *n* = 2 is less than half of its value when *n* = 3.

**Figure 5:**
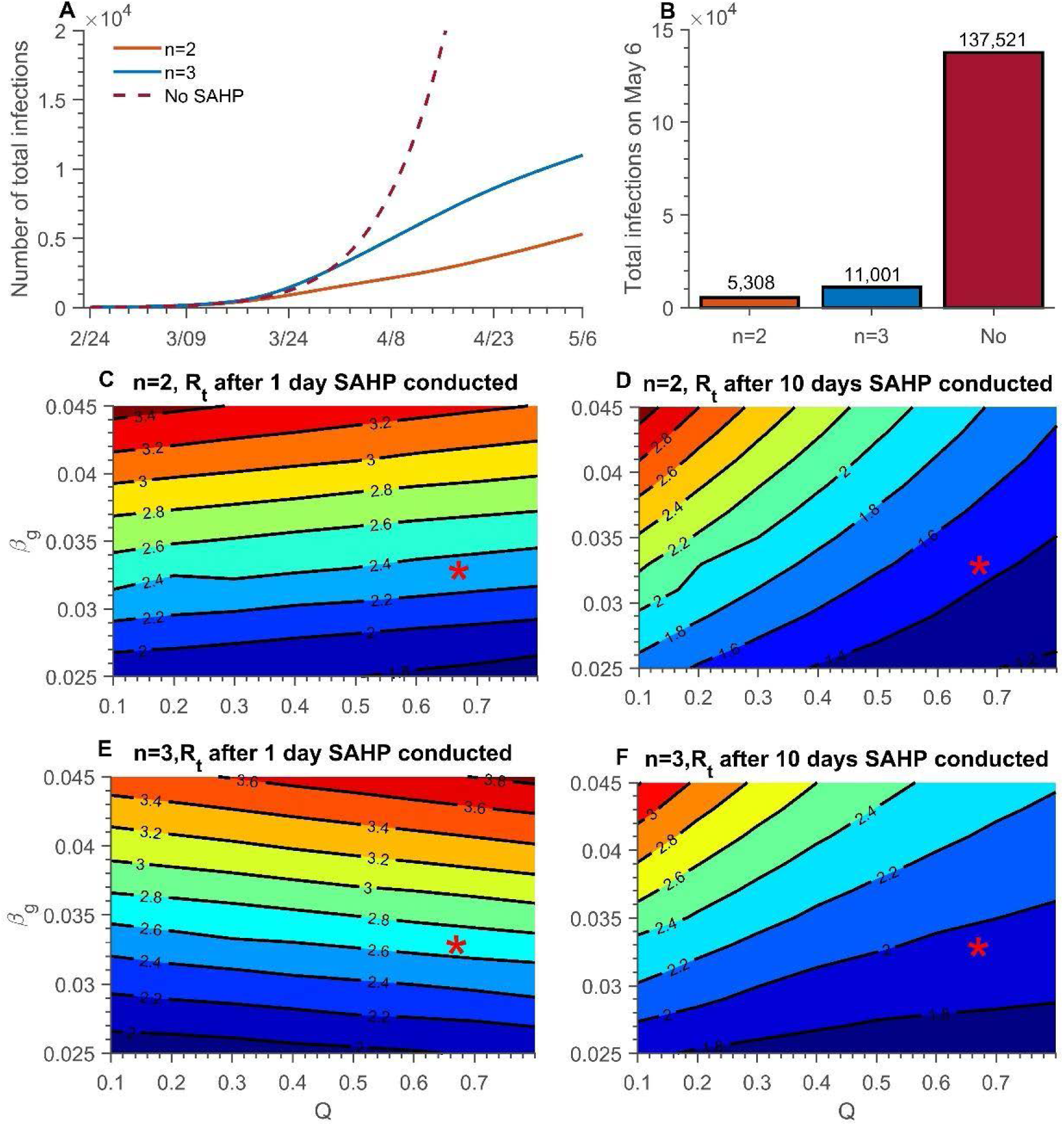
Effect of SAHP and different average household size. (A) The cumulative infection over time from Feb 24 to May 6 and (B) cumulative infection on May 6, without SAHP (dark red) and *n* = 2 (orange) and *n* = 3 (blue). Contour plot of *R*_*t*_ with different *β*_*g*_ and *Q* (C) one day after SAHP conducted, *n* = 2; (D) ten days after SAHP conducted, *n* = 2; (E) one day after SAHP conducted, *n* = 3; (F) after ten days after SAHP conducted, *n* = 3. *n* = average household size. *β*_*g*_ = probability of transmission per contact outside household. *Q* = maximum compliance rate.

However, in early phases of implementation of SAHP, due to the higher risk of transmission within the family, the number of infections was higher than when there was no SAHP (Figure 5A). This phenomenon is more pronounced when *n* is large (Figure 5E). With *n* = 2, *R*_*t*_ is seen to decrease as the maximum compliance rate (*Q*) increases for SAHP of both one- and ten-days duration (Figure 5C,D). In contrast, with *n* = 3, *R*_*t*_ is seen to decrease with *Q* for a 10-day SAHP but increase with *Q* after a SAHP of one day (Figure 5E,F).

The higher the value of *Q*, the sooner people comply with SAHP, which led to a lower total number of infections and deaths by May 6 (Figure 6A, B). If *Q* increases from 55% to 75%, the cumulative number of infections by May 6 will decrease by 63.2% (from 14032 to 5167), and the cumulative number of deaths will decline by 57.4% (from 504 to 215) when fixed Δ*T*_*Q*_ = 9 (Figure 6A). Furthermore, the reduction in contact rate is estimated to be 14%, declining from 7.7 to 6.6 (Figure 6C). If Δ*T*_*Q*_ is shortened from 9 days to 3 days and *Q*=0.65, the cumulative number of infections and cumulative deaths by May 6 will be reduced by 50.5% and 45.6%, respectively (Figure 6B). However, whether the epidemic continues to be controlled, or resurges, depends on the sustained compliance rate of SAHP.

**Figure 6:**
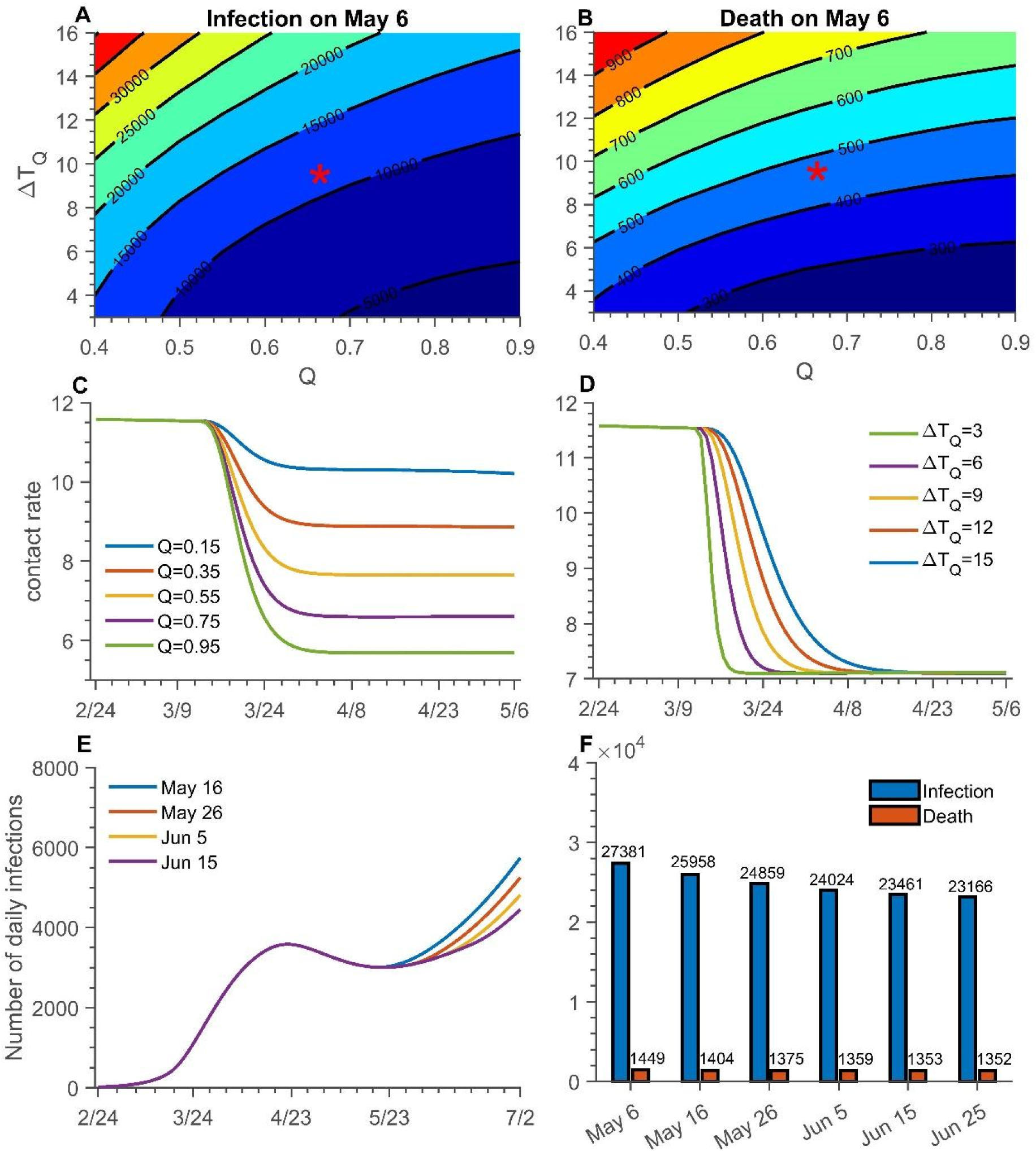
Effect of SAHP with maximum compliance rate, average completing time and length of SAHP. Contour plot of cumulative infection (A) and death (B) on May 6 with different Δ*T*_*Q*_ and *Q*. Red star represents the parameter values estimated from data and are used in simulations C, D, E, F. The contact rate change over time (C) under different *Q* with Δ*T*_*Q*_ = 9 and (D) under different Δ*T*_*Q*_with *Q* = 0.65. (E) The number of daily infection and (F) cumulative number of infections (blue bar) and deaths (orange bar) on Jul 2 with different length of SAHP, 55days (May 6), 65 days (May 16, blue), 75 days (May 26, orange), 85 days (Jun 5, yellow), 95 days (Jun 15, purple), 105 days (Jun 25). *Q*= maximum compliance rate. Δ*T*_*Q*_ = average completion time.

The population-wide governmental SAHP reduces the average contact rate outside the household, which affects the development of the epidemic. The compliance rate *Q* determines the extent to which the final contact rate can be reduced, while Δ*T*_*Q*_ determines the decrease in the contact rate (Figure 6C, D). If *Q* is increased to 95%, the contact rate will drop to 5.6.

The length of the SAHP conducted will affect the epidemic to a certain extent. The effect of an extended SAHP is not apparent. When the duration of SAHP is increased from 65 days to 95 days, the cumulative number of infections and cumulative deaths by July 2 only decreased by 9.6% and 3.6%, respectively (Figure 6F).

### 3.4 Threshold of contact rate and safe reopening

After the city’s reopening, the main factors affecting the epidemic are the contact (*β*_*g*_), the rate of detection of cases (and by consequence tracing and quarantine of contacts). When the symptomatic diagnosis’ completion ratio *d*_*c*_(*t*) is 97% (40%) (Figure 7B and 7A), if the contact rate is maintained at 11.58, *β*_*g*_ needs to be reduced by 5% (26%) to avoid epidemic resurgence; and if *β*_*g*_ is maintained at 1.9%, the contact rate needs to be reduced to 11 (9) (Figure 7A, B). When *d*_*c*_(*t*) is high and *β*_*g*_ =1.9% (the current state), *R*_*reopen*_ = 1.04, hence the city still face the risk of epidemic resurgence as the city reopens completely (Figure 7D, E). After reopening, if *β*_*g*_ increases to 2.2%, *R*_*reopen*_ = 1.2, there is a resurgence, while if *β*_*g*_ declines to 1.6%, *R*_*reopen*_ = 0.87, then reopening is safe, which are also shown in Figure 7E. Personal protection, stricter social distancing and other NPIs must be strengthened to maintain low risk as Toronto goes beyond stage 3 reopening.

**Figure 7:**
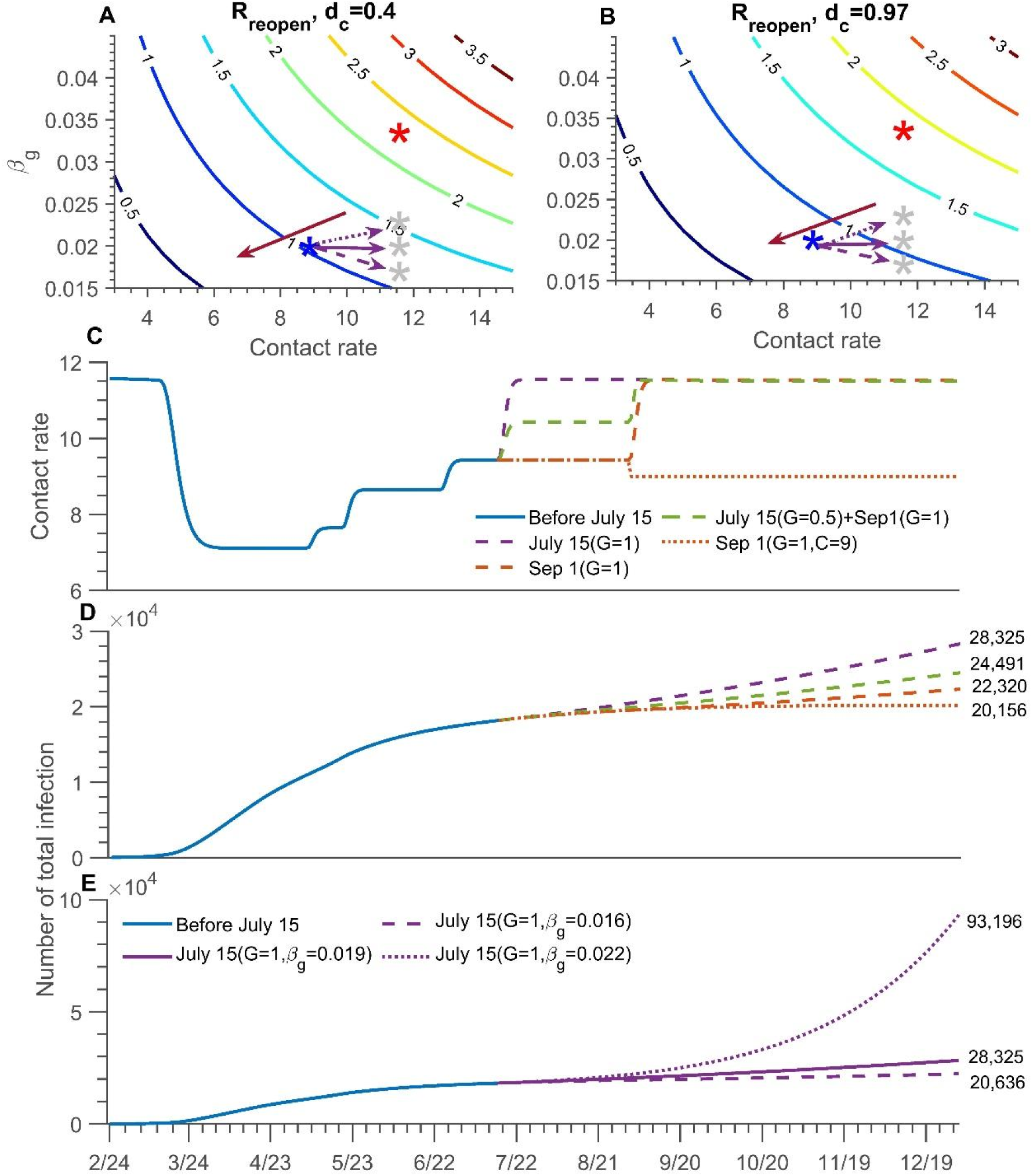
The risk of reopening, and different reopening scenarios. Contour plot of *R*_*out*_ with different *β*_*g*_ and C (A) *d*_*c*_= 0.4; (B) *d*_*c*_= 0.97. The red star is the initial status of *β*_*g*_ and C, and the blue is the current state. The grey star is the possible state after completely reopen in simulations E. The red arrow shows the low-risk direction with the safe reopening. (C) The change of contact rate and (D) the cumulative infection over time with different ways to reopen, fully reopening on July 15 (purple dash line), partially reopening on July 15 and then fully reopening on September 1 (green dash line), fully reopening on September 1 (orange dash line), fully reopening on September 1 and maintain contact rate is 9 (orange dot line). (E) The number of cumulative infections over time with *β*_*g*_ = 0.016 (dash line), 0.019 (solid line, current state), 0.022 (dot line) when fully reopening on July 15. *R*_*reopen*_ = the transmission risk after fully reopening. *d*_*c*_= completion ratio of symptomatic diagnosis. *β*_*g*_ = probability of transmission per contact outside the household. *C* =contact rate. *G* = going out rate.

Based on the current epidemic situation, combined with Toronto’s restart plan^7^, we projected the future trend of the epidemic and estimated risk presented by schools and workplaces reopening on September 1. We considered three different schools and offices reopening scenarios while keeping the current relevant parameters unchanged: (1) fully reopening on July 15, (2) partially reopening (50%) on July 15 and then fully reopening on September 1, (3) fully reopening on September 1. Compared to reopening on July 15, the risk of the epidemic resurgence is smaller if schools and workplaces fully reopen on September 1. Compared with the cumulative number of infections on July 15 (18,180), the cumulative increase in the number of infections in the three scenarios by the end of this year is expected to be 55.8%, 34.7% and 22.8%, respectively (Figure 7D). The corresponding change in contact rate is shown in Figure 7C. To ensure that the school reopening on September 1 does not cause resurgence, the contact still should be reduced. We show that there is a safe reopening of public places when reducing the contact rate to nine and maintaining current strict social distancing (Figure 7D).

## 4. Discussion

Using our novel model with household structure, we analyze the effect of the SAHP on the transmission of the COVID-19 using Toronto as a case study. SAHP has helped to control the epidemic and prevent the collapse of the healthcare system. However, in cities, such as Wuhan (China), SAHP was not effective at the early stage of the lockdown. This phenomenon can be related to average household size of 3.5 in Wuhan^24^, larger than the size 2.4 of Toronto^16^. Indeed, our results show that the smaller the average family size, the more obvious the mitigation effect. Therefore, the implementation of SAHP needs to be adapted to local conditions. For areas with large average family size, additional measures, such as the establishment of temporary shelter hospitals may be needed to reduce transmission in the home^3^.

Based on recent epidemic data of Toronto, simulations of different reopening strategies show the existence of risks in reopening schools and offices in September. If the daily per-capita contact rate is controlled at less than nine, the epidemic did not resurge in our simulations. Our study suggests that a gradual opening policy, for example, opening schools of different levels and offices at different times, would be safer. It is worth noting that this is based on the conclusions of the current epidemic development and control measures, and may not hold if public compliance deteriorates. Therefore, to better prevent and control the infection and restore economic activity, public health organizations need to continue to rigorous use of self-protective measures.

A lower probability of transmission when contacts occur, is provided by the use of mask, glove and facial shield usage, hand washing, disinfectants usages^25^. Recently, Canadian public health and government organizations have strongly recommended, and in some cases implemented mandatory use of masks during the epidemic, particular in indoor public places^26^. Maintaining social distance reduces the possibility of contact with the infected person, thereby the risk of infection. The combination of these measures made a significant contribution to the current epidemic control in Toronto, and, after reopening, it is important to pay attention on the conditions needed to relax these restrictions to avoid a new escalation of transmission.

Although the basic reproduction number (*R*_0_) is a key indicator of transmission, its estimation is not always feasible. Since our model includes asymptomatic cases, the *R*_0_ estimated based on the model (*R*_0_=2.36, 2.28-2.45) is higher than the estimate derived by case data (*R*_0_=1.45, 1.43-1.48). The instantaneous reproduction number *R*_*t*_ fluctuated with the SAHP in Toronto. However, after Toronto reopened into the first phase on May 19^7^, *R*_*t*_ gradually declined, possibly due to the strengthening of government regulations on personal protections’ use^7^. Although the contact rate may increase after reopening, the enhancement of personal protection is expected to reduce the probability of infection per contact (*β*_*g*_), thereby reducing the risk of the epidemic rebounding.

Using reported data to estimate the parameters of the model is widely questioned due to the time lag between the date of report and actual infection. However, these data show the efficiency of testing to a certain extent. In this study, we constructed the symptomatic diagnosis’ completion ratio (*d*_*c*_) as a model parameter which provides a new way of rethinking the use of report data and episode data together. The curve representing the ratio (Figure 1B) shows an increasing trend over time, indicating how public health’s response became more efficient as the pandemic grew. We also observe that *d*_*c*_ affects the achievement of *R*_*reopen*_ < 1. Less testing leads to new thresholds of the transmission rate and average number of contacts needed to achieve *R*_*reopen*_ < 1. Indeed, with a smaller *d*_*c*_, public health will need to strengthen NPIs and decrease the number of contacts. All these results confirm that factors such as the testing process, contacts and transmission play a crucial role in reducing the spread. Since relaxing one of them affects the others, it is important to take all into consideration when making a decision.

In conclusion, we explore the effect of SAHP by incorporating household structure and NPIs on the COVID-19 epidemic. Our findings highlight the contribution of current actions, such as school and workplace closures, revocation of gathering and public events, and stay-at-home measures, on mitigating the epidemic using Toronto as an example. The epidemic can be controlled if all the measures are strengthened simultaneously. The effect of SAHP has been almost wholly manifested after two months from its implementation. If the period of SAHP is extended, the impact on mitigating becomes not evident. Hence, this policy may be relaxed when the epidemic is effectively alleviated, then combined with social distancing, wearing PPEs, increasing the detection and isolation rate of symptomatic infections (with associated contact tracing and quarantine), to maintain control of the epidemic and reduce the burden on the healthcare system. We then establish that a safe and full reopening of all activities may be possible, if citizens strictly adhere to correct and persistent use of personal distancing and transmission prevention measures.

## Data Availability

All data used in this study are from public available source for Ontario and Toronto. The links are given below.

https://www.toronto.ca/home/covid-19/

https://www12.statcan.gc.ca/census-recensement/2016/dp-pd/prof/details/Page.cfm?Lang=E&Geo1=CSD&Code1=3520005&Geo2=PR&Data=Count&B1=All

## Author Contributions

Research design: H.Z., P.Y., JuanL, N.O., J.B., J.H.; Literature search: Q.L., E.A., P.Y., JuanL, T.Z., Y.T.; Data collection: T.Z., Q.L., Y.T., P.Y., JuanL.; Modeling: H.Z. and all; Model analysis: JunL., P.Y., JuanL., E.A., H.Z.; Simulations: P.Y., JuanL., E.A., T.Z., Y.T.,JunL; Draft preparation: P.Y., JuanL, E.A., Q.L., T.Z., Y.T., H.Z.; Writing-reviewing-editing: H.Z.,N.O, B.S., J.H., EvgeniaG., EffieG., S.C., J.A., J.B., J.W., J.D. K., I.M., Supervision: H.Z.

## Appendix A

### Modeling

#### 1. Description

This study considers the entire population of Toronto with the “stay at home” policy (SAHP) that was enacted on March 12th and gradually relaxed after May 6^7^, as well as the document “A Framework for Reopening our Province” Ontario released on April 27^30^. The province will gradually reopen all workplaces and public spaces. Stage 1, which began on May 19, allowed the opening of select workplaces and some small gatherings. On Jun 24, the city of Toronto enters Stage 2 of reopening, opening more workplaces and outdoor spaces, allowing gatherings of up to 10 people^7^. We divide the population into two groups: one consisting of individuals who follow SAHP (marked by subscript *q*) and another consisting of individuals who do not opt for this intervention (marked by subscript *g*). Due to influences of self-protection consciousness and severity of the epidemic, people are assumed to move from one group to another with stay-at-home rate (denoted by *q*(*t*)) or going-out rate (denoted by *g*(*t*)). We note that we omit demographic components, such as immigration, birth and natural death.

A detailed description of dynamical transmission of COVID-19 is described in the flowchart (Fig. 3). Let *N*_*i*_(*t*) (*C* = *g, q*) be the total number of individuals in each sub-group, *g, q*, at time t. Each subpopulation is further the divided into Susceptible (*S*_*i*_(*t*)), Exposed (*E*_*i*_(*t*)), Asymptomatic (subclinical) infection (*A*_*i*_(*t*)), Infectious pre-symptomatic (will eventually show symptoms) (*I*_*i*1_(*t*)) and Infectious symptomatic (*I*_*i*2_(*t*)). Both *A*_*i*_(*t*) and *I*_*i*1_(*t*) are considered to be infectious virus carriers. We assume that individuals in *A*_*i*_(*t*) will never show symptoms, while individuals in *I*_*i*1_(*t*) develop into symptomatic classes (*I*_*i*2_(*t*)) after a specified period of time. Mild symptomatic infections in classes (*I*_*i*2_(*t*)), may choose to either isolate themselves at home (or other places). If the quarantine is respected well enough, these infections will be fully isolated and, consequently, will not contribute to the spread of the virus. Otherwise, they are still a source of infection until recovery.

As the disease progresses, some mild infections may become severe and require hospitalization. We include two further compartments: the fully isolated (*W*(*t*)), and the hospitalized (H(t)) who are all severely affected. It is assumed that neither of these compartments contribute to infection transmission. Through a numerical analysis of H(t) and W(t) relevant parameters, we will present a pre-estimation of the ratio of mild to severe infections during the epidemic. We will also explore the influences of some measures (such as hospital capacity, testing and isolation) on the development of the disease.

Based on the classical SEIR framework, a household-based transmission model will be proposed to describe the impact of SAHP on the development of the epidemic. Considering that an infected person quarantined at home is interacting only with family members, the number of contacts is limited, so we will use the standard incidence rate in modelling.

Although home transmission is relatively strong, it only involves limited family members. To reflect this, and capture disease transmission within families, we separate people who follow the SAHP into households.

For family members following SAHP, susceptible individuals (*S*_*q*_(*t*)) will only be infected by infectious individuals in the home *A*_*q*_(*t*), *I*_*q*1_(*t*) *or I*_*q*2_(*t*). When no cases are reported in a household, the family will be safe and will no longer be involved in the transmission of COVID-19. Additionally, infections who are completely isolated will not be involved in transmission.

#### 2. Rates definition

Next, we will present the dynamical models for SAHP non-compliant, SAHP compliant and isolation population, respectively. First, we will describe the key rates on which the model is based.

##### Stay-at-home rate

Before the government implemented SAHP on March 12^7^, due to the impact of self-prevention awareness and the severity of the epidemic, a small number of people would consciously stay at home, so we assume that the stay-at-home rate is a very small constant, which we express as

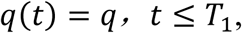

where *T*_1_ is the time when the SAHP is implemented, and *q* is the average daily stay-at-home rate before the policy is put into action.

After the SAHP was implemented, some people chose to stay at home based on their own behaviors and their knowledge of the epidemic. We denote the maximum compliance rate (*Q*_1_) as the maximum proportion of the number of people in the group that will carry out SAHP, which is used to reflect the degree of the behavioral tendency of the population to change their original daily lifestyle and accept the SAHP under the requirements of prevention and control policies after the outbreak. The implementation of SAHP will directly affect the stay-at-home rate

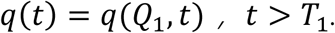

Then we have

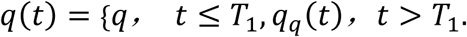

We assume that τ is a random variable which describes how long it will take the five groups *S*_*g*_, *E*_*g*_, *A*_*g*_, *I*_*g*1_, *I*_*g*2_ to complete the stay-at-home process when conducting SAHP. Although *I*_*g*2_ is the symptomatic compartment, it should be the same as the other four categories before tested and confirmed. Hence, τ follows a Gamma distribution

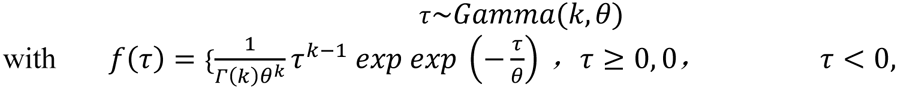

where 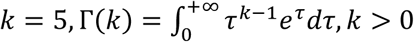.

The expectation of *τ* = *E*(*τ*) = *k* * *θ* = Δ*T*_*Q*_ (Δ*T*_*Q*_ is the average completing time for all those who conducted the SAHP), and *f*(*τ*) is the probability that those in the five groups will accomplish stay-at-home process in *τ* days.

The total population that may conduct SAHP of Toronto at *T*_1_ is *P*_1_ = *S*_*g*_(*T*_1_) + *A*_*g*_(*T*_1_) + *E*_*g*_(*T*_1_) + *I*_*g*1_(*T*_1_) + *I*_*g*2_(*T*_1_). The number of people who accomplished stay-at-home process on *T*_1_ + *τ* days was Δ*P*_1_(*T*_1_ + *τ*) = *Q*_1_ * *P*_1_ * *f* (*τ*) = *Q*_1_ * *f*(*τ*) * *P*_1_. Let *Q*(*T*_1_ + *τ*) be the daily stay-at-home rate on day *T*_1_ + *τ*, then *Q*_1_(*T*_1_ + *τ*) = *Q*_1_ * *f* (*τ*). And it satisfies 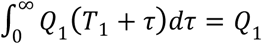.

We also assume that each group has the same daily stay-at-home ratio, *q*_*q*_(*T*_1_ + *τ*), which is the daily stay-at-home rate of the people who began to stay at home on day *T*_1_ + *τ*. Then the number of people newly stay-at-home on that day is

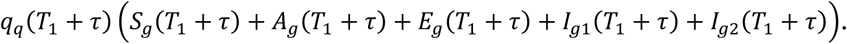

The newly stay-at-home number on day *T*_1_ + *τ* is equal to the number of people conducting SAHP on that day, i.e.,

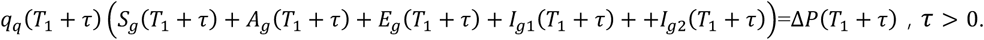

Hence, we have

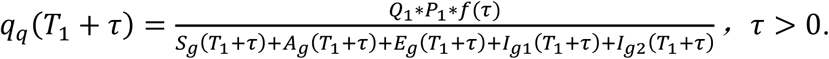

Let *t = T*_1_ + *τ*, then

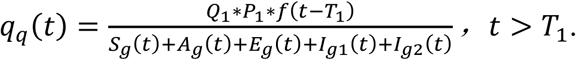

According to the relative policies of Toronto, people who are detected to be COVID-19 positive need to stay at home and self-isolate for 14 days^15^.Combined with the flowchart shown in Fig.3, there are three different ways to allocate infectious patients: to be hospitalized, to isolate at home, or to isolate in a place other than home. Due to the strengthening effect of testing, the stay-at-home rate of the infected cases with symptoms is much higher than others. Here, we modify the quarantined rate of *I*_*g*2_ (separately rewritten as *q*_*g*2_(*t*)) to be *q*_*g*2_(*t*) = *q*(*t*) + *εd*_*c*_ (*t*), where *d*_*c*_(*t*) is the completion rate of diagnosis of all symptomatic infections. *d*_*c*_(*t*) obtained from the onset data and the reported data shown in Section 2, and *ε* is an adjustment parameter to describe the impact of testing on the quarantine rate of *I*_*g*2_. Here, it is assumed that *q*_*g*2_(*t*) = *q*(*t*) if there is no testing.

##### Going out rate

Let*T*_2_ (*T*_2_ > *T*_1_) be the day on which the SAHP is announced to be relaxed. That is, some people would be encouraged to go outside home after that day. Similar to the formula design process of *q*(*t*), we now determine *g*(*t*), the proportion of households that are not stay-at-home versus all households, which is given by

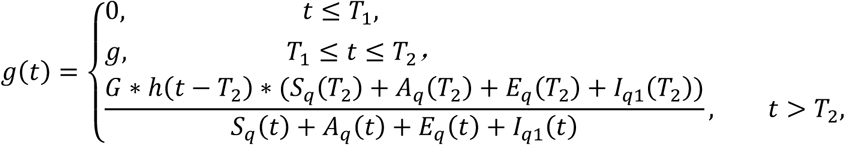

where *g* is a small positive constant, *G* is the maximum proportion of the population who will not continue to stay at home compared to the total size of the stay-at-home population at time *T*_2_,

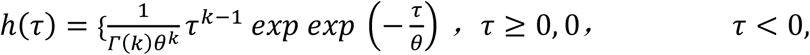

*k* = 4 and *τ* = Δ*T*_*G*_, where Δ*T*_*G*_ is the average completion time for all people who stay at home (except those with symptoms) to go outside.

#### 3. Models

##### Stay-at-home and Isolation

According to the infection and development process of the disease in the human body, at time t, an individual in a household can belong to one of the following categories: *S*_*q*_(*t*), *E*_*q*_(*t*), *A*_*q*_(*t*), *I*_*q*1_(*t*), *I*_*q*2_(*t*), *H*_*q*_(*t*) or *W*_*q*_(*t*), or may be recovered, Corresponding to each disease class, we assign the number of individuals in each household to be *i, j, k, l, m, x, y, z*, respectively, and limit households to a size of *n* such that *n* = *i* + *j* + *k* + *l* + *m* + *x* + *y* + *z*. Therefore, each household at most consists of *n* different categories of individuals. Based on the classification and combination of individuals in households, all possible types of households in Toronto are 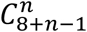.

For each household type, the dynamics are determined by eight processes: within-household transmission; disease progression from Exposed to Asymptomatic infection or Infection without symptoms; disease progression from Infection without symptom to Infected with symptoms; recovery from Asymptomatic infection; recovery from Infected with symptoms; hospitalization of Infected with symptoms; isolation of Infected with symptoms; and newly entered stay-at-home. Then the variation of the number of households *P*_*i,j,k,l,m,x,y,z*_ with respect to time *t* can be given by

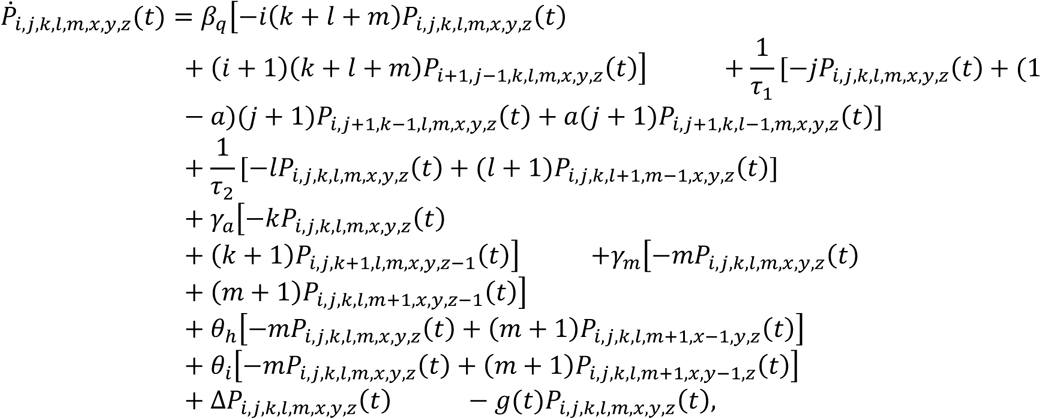

where *P*_*i,j,k,l,m,x,y,z*_ (*t*) ≥ 0 should be satisfied, *g*(*t*)*P*_*i,j,k,l,m,x,y,z*_(*t*) should be ignored for *m* ≠ 0, and Δ*P*_*i,j,k,l,m,x,y,z*_(*t*) is the number of new stay -at-hom e households with *i* susceptible, *j* exposed, *k* asymptomatic (subclinical) infection, *l* infectious without no symptoms, *m* infected with symptoms, *x* hospitalized, *y* isolated and *z* removed members,

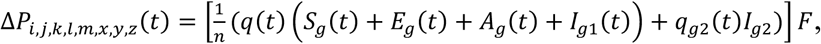

where [·] is an integral function to return the value of a number rounded downwards to the nearest integer and *F* i s the probability of each type of newly added quarantine household when *n* = 2,

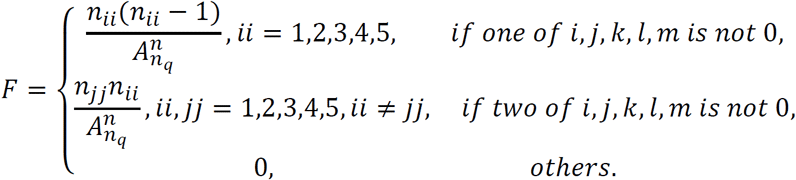

when *n* = 3,

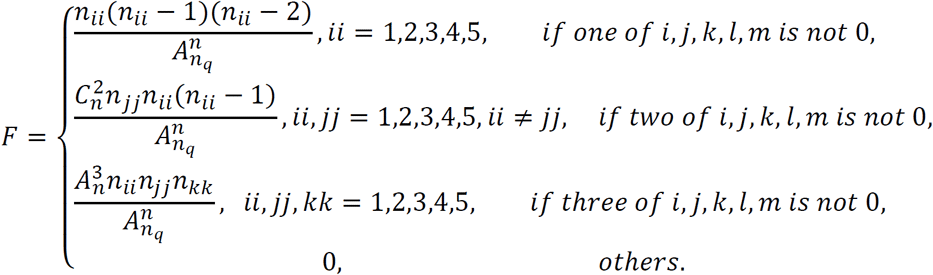

With 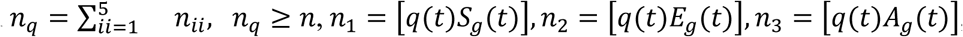, *n*_4_ = [*q*(*t*)*I*_*g*1_ (*t*)], *n*_5_ = [*q*_*g*2_(*t*)*I*_*g*2_].

With the above, we have the model describing the dynamics of the groups with stay-at-home and isolation as

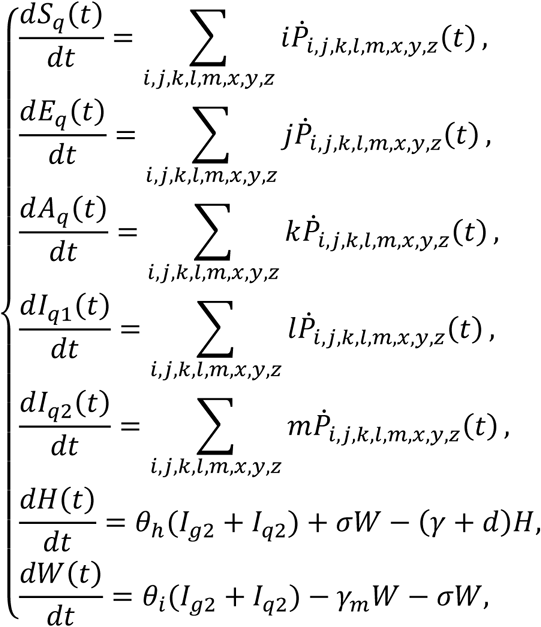

where all parameters are positive, the interpretation of the variables and parameters are summarized in Table 2 and 3.

##### SAHP non-compliant population

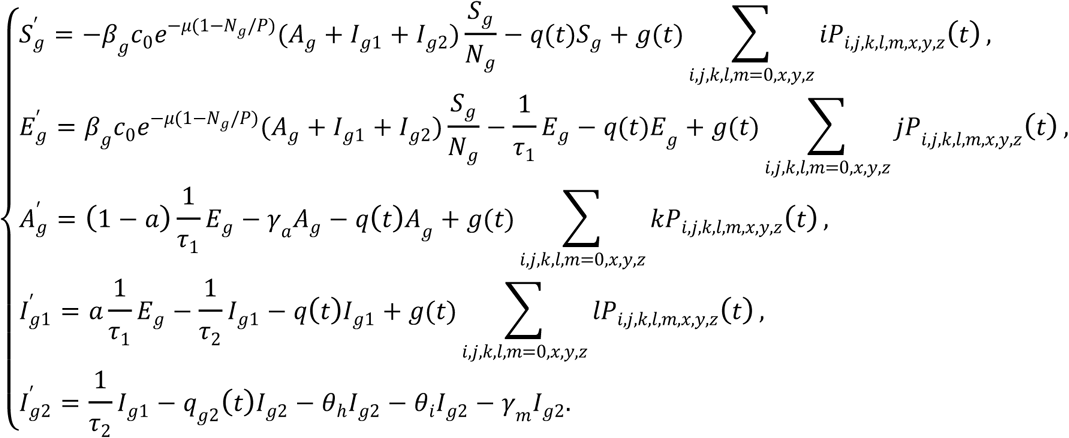

where *N*_*g*_(*t*) = *S*_*g*_(*t*) + *A*_*g*_(*t*) + *E*_*g*_(*t*) + *I*_*g*1_(*t*) + *I*_*g*2_(*t*), *P*_*i, j,k, l, m, x, y, z*_ is the number of households with *i* susceptible, *j* exposed, k asymptomatic (subclinical) infection, l infectious without no symptoms, m infected with symptoms, x hospitalized, y isolated and z recovered members. The contact rate is 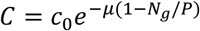. all parameters are positive, and the interpretation of other variables and parameters are given in Tables 2 and 3.

